# Intracranial Biomarkers for Anterior Thalamic Deep Brain Stimulation in Epilepsy: a long-term observational study

**DOI:** 10.1101/2025.09.12.25335307

**Authors:** Giovanna Aiello, Lennart Stieglitz, Anina Belvedere, Tena Dubcek, Rafael Polania, Lukas Imbach

**Author notes:** Corresponding author. (L.I.). These authors contributed equally to this work as co-senior authors (Rafael Polania, Lukas Imbach). (G.A) (R.P).

## Abstract

Deep brain stimulation (DBS) of the anterior nucleus of the thalamus (ANT) is an established treatment for patients with drug-refractory epilepsy (DRE), yet long-term therapeutic outcomes are highly variable and challenging to predict. This variability is compounded by the delayed and gradual effects of DBS, the difficulty of consistent seizure monitoring, and the absence of physiological biomarkers to inform treatment. In this study, we analyzed longitudinal intracranial recordings over a four-year observational period from a cohort of 22 patients with ANT-DBS. Our primary goal is the identification of neurophysiological signatures that could predict and track clinical DBS response.

Our results show that DBS-responders and non-responders exhibit distinct ANT spectral trajectories over time, with responders showing a progressive increase in higher frequencies (*β*_1_, *γ*) and decreased lower-frequency (*δ, θ*) activity, as compared to non-responders. Notably, these dynamic biomarkers, particularly high-to-low frequency ratios (e.g., *β*_1_/*θ*), enabled the early discrimination of clinical outcomes. Additionally, we provide evidence of robust circadian and multidien rhythms in ANT local field potentials, with further analyses supporting the feasibility of adaptive stimulation protocols to improve therapeutic outcomes.

Together, these findings propose ANT spectral dynamics in outpatient settings as a promising tool for early prediction of therapeutic efficacy and pave the way for biomarker-guided optimization of DBS therapy in epilepsy.

**Highlights:**

- Longitudinal analysis of spectral activity evolution in the anterior thalamic nucleus (ANT) revealed different temporal dynamics in responders and non-responders to ANT-Deep Brain Stimulation (DBS).
- High-to-low frequency power ratios (e.g., *β*_1_/*θ*) enable discrimination between super-responders and non-responders for assessing therapeutic efficacy, offering a potential tool for early therapy evaluation.
- ANT activity exhibits circadian and multidien fluctuations, reinforcing its potential as a signal source for adaptive DBS strategies tailored to individual brain states.

## Introduction

Drug-refractory epilepsy (DRE) affects approximately one-third of epilepsy patients ^1(p20),2^. For those not eligible for resective epilepsy surgery, due to multifocal or generalized seizure onset, deep brain stimulation (DBS) is an alternative treatment option^3–5^ whose safety and efficacy were validated in a double-blind, randomized, multi-center controlled clinical trial (SANTE trial)^6^.

However, we still observe a large variability in responsiveness to anterior thalamic (ANT) DBS^7^. For instance, median seizure reduction at 5 years post-implantation is reported at 65% in the SANTE trial^8^ and 56% in the recently published MORE registry^9^, while seizure freedom is only achieved in 16%^10^ of patients^11,12^. As the accepted concept of ‘DBS responsiveness’ only applies to patients who experience a reduction in seizure frequency of at least 50%, optimal treatment strategies should aim to maximize seizure reduction rates in order to significantly improve quality of life.

A growing body of evidence suggests that DBS acts by chronically modulating epileptic networks rather than working as an acute seizure-suppressor^13–16^. The most compelling support for this hypothesis is the long-term improvement of patients, who undergo a gradual reduction of seizures within a time scale of years^6,8,11,17,18^. In the SANTE trial, the median seizure reduction increased up to 75% at 7 years follow-up. This strongly supports the idea that thalamic neuromodulation involves neuronal reorganization and engages neuroplastic mechanisms, e.g. long-term potentiation or altered gene expression^19,20^. These findings have also sparked a growing interest in identifying neurophysiological signatures that could predict the long-term response to treatment by reflecting favorable adaptations in brain networks. On the other hand, these slow-evolving changes make it more difficult to evaluate therapeutic effectiveness in the early stages and to decide whether treatment adjustments or re-evaluation are needed.

Alongside the neural changes driven by chronic neuroplasticity, implanted patients also undergo multiple device-tuning sessions to identify the optimal combination of stimulation parameters that can maximize seizure reduction. Nevertheless, stimulation protocols are often adjusted through trial-and-error approaches due to the lack of standardized guidelines for parameter optimization. This process is further complicated by the fact that approximately one-third of patients fail to consistently report seizures — typically the primary clinical target variable — with studies indicating that nearly 50% of seizures go undocumented^21–23^. Finally, reliable assessment of seizure frequency reduction requires monitoring over weeks or even months, potentially delaying meaningful improvements^24^ because the therapeutic effects of stimulation emerge gradually.

For these reasons, there is an urgent need for a framework to define biomarkers that correlate with the wide variability in patient responsiveness to optimize therapeutic outcomes on an individual basis. This challenge involves multiple dimensions, including (i) determining whether a patient’s neurophysiological profile influences their likelihood of responding to therapy^25^ (predictive biomarkers) and (ii) identifying robust quantitative indicators that track treatment efficacy online.

Recent technological and conceptual advancements in DBS have opened new avenues towards the concept of therapy personalization. In particular, the emergence of stimulation devices able to both record and stimulate (i.e., bidirectional) offers the possibility to chronically monitor neural activity and adapt stimulation based on real-time or long-term brain dynamics. These tools have fueled interest in defining neurophysiological biomarkers that can optimize therapy in a data-driven, patient-specific manner^26–31^. Furthermore, the ability to record from the implanted device enables continuous data collection outside of the clinical setting.

While these biomarkers may aid patient selection and capture the slow and progressive modulation of brain networks over long time periods, another aspect of personalizing DBS involves the faster, cyclical fluctuations in brain excitability that influence seizure likelihood. Seizure risk is not constant but varies with intrinsic biological and physiological rhythms, including circadian (24-hour) ^32–34^, multidien, seasonal, and menstrual cycles. These rhythms influence seizure timing and severity^35–41^ and are linked to seizure clustering^32,36,42–44^. In addition to these predictable cycles, growing evidence supports the existence of pro-ictal states—fluctuations in brain states that increase seizure propensity^45,46^—and their potential impact on stimulation efficacy.

Importantly, aligning neuromodulation protocols with such temporal dynamics could enhance therapeutic outcomes through state-dependent stimulation strategies^47 15 48 49^. However, it remains to be elucidated whether ANT activity can reliably detect such states and provide meaningful information for adaptive stimulation strategies.

Here, we investigate how ANT activity-based biomarkers can serve three distinct purposes. First, anterior thalamic local field potentials (ANT-LFP) evolve alongside treatment progression, offering a reliable quantitative framework of chronic neuromodulation. Second, their long-term correlation with therapeutic efficacy may enable early prediction of clinical DBS outcomes. Third, the presence of specific neurophysiological oscillations can be utilized to inform state-dependent neuromodulation therapies.

## Materials and Methods

### Patient Population and Surgical Implantation

A cohort of 22 patients (10 females) diagnosed with drug-resistant focal epilepsy (DRE), with an average age of onset of 16.27 ± 14.47 years underwent bilateral implantation of deep brain stimulation (DBS) leads in the anterior nucleus of the thalamus (ANT) (Table 1). Average age at implantation was 37.59 ± 12.12 years. All surgeries were conducted between 2018 and 2024 at the Department of Neurosurgery, University Hospital Zurich by stereotactic implantation with the *neuromate®* robotic system (Renishaw Mayfield SA). Detailed information about the pre-surgical evaluation and surgical implantation procedures is available in our previously published work^50^. Informed consent was obtained from all patients or their legal representatives in compliance with the Declaration of Helsinki, and the study was approved by the local ethics committee (Kantonale Ethikkommission, KEK Zurich). However, this consent did not include provisions for making individual data publicly available. Treatment response was evaluated based on self-reported seizure frequency at last follow-up.

**Table 1.** Patient Demographics. Patient demographics (including gender, age of onset, age at implantation, seizure onset lobes, Engel/ILAE at last follow-up, responsiveness, types of recordings) is reported. Ages are reported as non-overlapping age ranges. **Abbreviations** *Lobes included in the seizure onset* (F = frontal, P = parietal, T = temporal, O = occipital, 1= left, 2 = right) *Types of recording* (LS = LFP Streaming, CS= Chronic Sensing); “t=I” surgery date (implantation); “t=PreI” : pre-implantation; “t=end” : last clinical follow-up. Does not necessarily include a recording.; “PI”= post-implantation. Response is indicated as one of four levels (0=0-25%, 1=25-50%, 2=50-75%, 3>75%). Percentages indicate seizure frequency reduction compared to pre-implantation baseline).

| ID/Gender | Age / Age of Onset | Age at $t = I$ | $ASM_{t=PreI}$ / $ASM_{t=end}$ | Lobes in SOZ | Engel /ILAE ($t = end$) | Response ($t = end$) | Type of recordings (LS/CS) | First recording PI (days) | Last recording PI (days) |
| --- | --- | --- | --- | --- | --- | --- | --- | --- | --- |
| *S1/M* | 31-35/<br>1-5 | 21-25 | 2/2 | T1, T2, F2 | 4/5 | 0 | LS | 70 | 533 |
| *S2/M* | 46-50/<br>21-25 | 36-40 | 3/3 | T1 T2 | 3A/4 | 2 | LS/CS | 97 | 944 |
| *S3/M* | 31-35/<br>1-5 | 26-30 | 4/6 | T1 T2 F1 F2 | 4B/5 | 1 | LS/CS | 270 | 1452 |
| *S4/W* | 71-75/<br>61-65 | 66-70 | 3/2 | T1 T2 P1 P2 | 1A/1 | 3 | LS | 260 | 260 |
| *S5/W* | 51-55/<br>16-20 | 46-50 | 4/4 | other | 3A/4 | 3 | LS/CS | 43 | 844 |
| *S6/W* | 56-60/<br>16-20 | 51-55 | 2/2 | T1, T2 | 2B/3 | 3 | LS | 4 | 901 |
| *S7/M* | 46-50/<br>6-10 | 41-45 | 4/4 | F1 F2 | 3A/4 | 2 | LS/CS | 112 | 1092 |
| *S8/W* | 26-30/<br>1-5 | 26-30 | 5/2 | F1 | 4B/5 | 1 | LS | 63 | 594 |
| *S9/W* | 56-60/<br>1-5 | 51-55 | 6/6 | T1 T2 F1 F2 | 3A/4 | 1 | LS | 1 | 742 |
| *S10/W* | 51-55/<br>16-20 | 46-50 | 2/2 | T1 T2 P2 F2 | 2B/3 | 3 | LS | 64 | 147 |
| *S11/M* | 46-50/<br>11-15 | 41-45 | 5/5 | F1 F2 | 3A/4 | 2 | LS/CS | 641 | 879 |
| *S12/M* | 36-40/<br>16-20 | 31-35 | 4/4 | T1 T2 | 3A/4 | 2 | CS | 235 | 235 |
| *S13/M* | 26-30/<br>11-15 | 26-30 | 2/2 | T1 T2 | 3A/4 | 3 | LS/CS | 214 | 1286 |
| *S14/M* | 51-55/<br>41-45 | 51-55 | 3/2 | T1 F1 F2 | 3B/4 | 3 | LS/CS | 110 | 831 |
| *S15/W* | 31-35/<br>16-20 | 26-30 | 6/4 | T2 P2 O2 | 3A/4 | 2 | LS | 1 | 418 |
| *S16/W* | 26-30/<br>6-10 | 26-30 | 4/3 | F1 F2 | 4B/5 | 0 | LS | 76 | 600 |
| *S17/M* | 36-40/<br>1-5 | 36-40 | 3/2 | T1 T2 | 3A/4 | 2 | LS/CS | 7 | 706 |
| *S18/M* | 26-30/<br>26-30 | 26-30 | 3/2 | T2, AHR | 4B/5 | 1 | LS/CS | 6 | 484 |
| *S19/W* | 36-40/<br>11-15 | 36-40 | 2/1 | F1 F2 | 3A/4 | 2 | LS | 2 | 376 |
| *S20/M* | 26-30/<br>6-10 | 26-30 | 3/3 | T1 F1 | 3A/4 | 2 | LS | 8 | 231 |
| *S21/W* | 21-25/<br>16-20 | 16-20 | 4/3 | P1 P2 O1 T1 T2 | 3A/4 | 2 | CS | 14 | 14 |
| *S22/M* | 36-40/<br>16-20 | 36-40 | 4/4 | T1 T2 | 4/5 | 0 | LS/CS | 21 | 111 |

### Deep Brain Stimulation

All participants were implanted with quadripolar DBS leads (3389, Medtronic) connected to a Medtronic PERCEPT^™^ system. Chronic continuous (non-cycling) stimulation was administered, with a default stimulation frequency of 145 Hz. Stimulation parameters, including active contact, stimulation modality (monopolar or bipolar), and amplitude, were tailored individually based on periodic follow-ups and treatment response (Supplementary Figure 1). Here after, the four contacts of one DBS lead will be referred to as 0,1,2 and 3, with 0 being the most inferior contact on the lead, and 3 the most superior. Contacts are equally named on the left and the right lead.

### Data acquisition and selection

During regular out-patient follow-ups, patients underwent two types of recording to monitor treatment response:

1. *Local Field Potential (LFP) Streaming* During out-patients visits, we recorded LFP data from three non-adjacent contact pairs in each side (0-3, 1-3, 0-2), without ongoing stimulation (i.e., passive streaming mode), in standardized recording settings: patients were awake, relaxed, with eyes closed and in a sitting position. The average recording time was 75.73 ± 32.43 seconds. The raw signal (LFP in *µ*V) has a sampling frequency of 250 Hz.
2. *Chronic Sensing* In this modality, patients were monitored for a maximum consecutive duration of 60 days, gathering one data sample every 10 minutes (sampling frequency of 144 samples/day) in a predefined frequency band. Thus, each data point represents the sum band power in a specific frequency band, which was tailored on an individual basis based on the most prominent peak in the ANT-LFP periodogram. The non-adjustable band-width for this read-out was 5 Hz and the power was recorded from one pre-selected montage (bipolar), adjacent to the active (stimulation) contact^51^.

Patients were monitored in both modalities multiple times at different periods post-implantation (up to 4 years, Figure 1). LFP streaming data was gathered during all outpatient visits. Chronic sensing was only activated if the stimulation setting allowed for it and a spectral peak was visible in the raw LFP spectrum.

**Figure 1.**
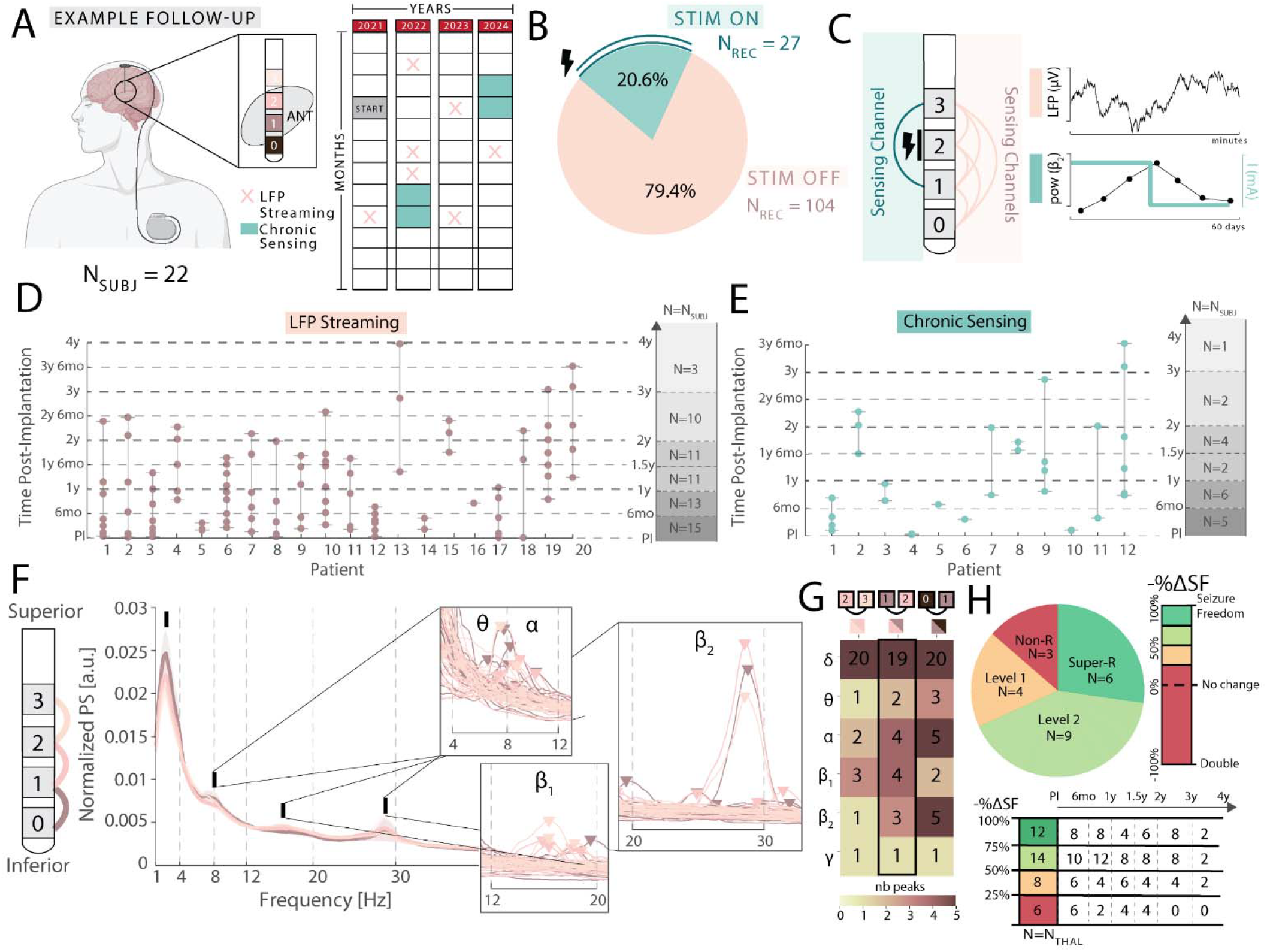
A) Schematic of the experimental design. Each of the N=22 patients implanted with DBS in the ANT was recorded throughout multiple years. “START” indicates the implantation date. B) Both recordings in-clinic (Local Field Potential Streaming) and Chronic Sensing were performed. C) B) Distribution of the available recordings and sensing configurations in the two modalities. C) For each patient recorded in the two modalities, distribution of follow-ups at time-post implantation is shown. F) For each bipolar montage (indicated with different shades of pink), the mean periodogram across subjects is represented (together with the standard error of the mean, as a shaded area). The relevant frequencies in the periodogram are indicated with bold black dashes and zoomed-in with the peak detections shown for *θ, α, β*_1_, *β*_2_. G) The number of subjects with a peak detected for each bipolar montage and each frequency is reported in the heatmap. H) Distribution of the four classes of responsiveness, together with their representation over time.

## Data processing and extracted features

### LFP Streaming

In the *LFP Streaming* modality, three voltage differences were measured at the contact pairs 0-3, 1-3 and 0-2 (here after, *V*_0|3_, *V*_1|3_, *V*_1|2_). We re-referenced to neighboring bipolar configurations by calculating the linear combinations of *V*_0|3_, *V*_1|3_, *V*_1|2_, for example: *V*_0|1_ = *V*_0_ − *V*_1_ = *V*_0_ − *V*_0_ − *V*_3_ − (*V*_1_ − *V*_3_) = *V*_0|3_ − *V*_1|3_ ^51,52^ Following the same procedure, we derived *V*_0|1_, *V*_1|2_ and *V*_2|3_. The re-referencing was performed to ensure constant distance between bipolar contacts to reduce the bias in amplitude baseline differences and to remove the influence of distance-based filtering of high or low-frequency components of the signal^53^. Extracting these three combinations allowed us to compare spectral properties of different locations within the ANT.

Each patient’s first measurement post-implantation (*t* = *t*_*0*_) was considered as the “post-implantation” baseline. We did not analyze pre-implantation ANT measurements via, for instance, stereoelectroencephalography (sEEG) in the pre-surgical evaluation phase^54,55^ or intraoperatively during the implantation.

### Power spectral analysis

We preprocessed the raw data to extract the power spectral density by firstly applying a Notch filter at 50 Hz and its harmonics, followed by a 4^th^ order Butterworth bandpass filter in the frequencies of 1 to 70 Hz. We then epoched the data in 4-seconds windows with no overlap^50^. Band-wise power spectrum was then calculated by averaging the power in six bands of interest: *δ* (1-4 Hz), *θ* (4-8 Hz), *α* (8-12 Hz), *β*_1_ (12-20 Hz), *β*_2_ (20-30 Hz), *γ* (30-70 Hz). To ensure across-subjects comparison, we normalized the power spectra to their maximum value.

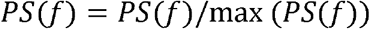

We firstly assessed the change over time of the analyzed bands by grouping the time post-implantation in 6 “intervals”: (i) *post-implantation to 6 months*, (ii) *from 6 months to 1 year*, (iii) *from 1 year to 1 year and a half*, (iv) *from 1 year and a half to 2 years*, (v) *from 2 to 3 years* and (vi) *from 3 to 4 years*. We then normalized the power spectrum of each frequency band (fb) by dividing it to the power spectrum on the first observation post-implantation, to assess its evolution for each thalamus j (T_j_) separately:

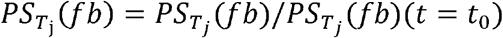

Each bipolar contact of interest (*V*_0|1_, *V*_1|2_ and *V*_2|3_) was analyzed separately to assess how location within the ANT affects neurophysiological DBS response.

### Chronic Sensing data

Data recorded in Chronic Sensing was under *continuous* stimulation and gathered from only one non-adjacent bipolar montage (surrounding the stimulating contact(s))^51^. As an example, if the stimulating contact is contact 2, the signal will be recorded in the contact pair 1-3 (*V*_1|3_). Based on the individual power spectrum of each subject, the most prominent peak in the ANT was detected, and its specific frequency (with a band-width of 5 Hz) was monitored continuously, sampling every 10 minutes during a maximum of 60 days^51,52^. We separately analyzed thalamic recordings from both sides. Moreover, the same patient might have more than one 60-days recording over time (although not necessarily consecutively collected).

To assess the presence of *ultradian, circadian and multidien* rhythms, we first pre-processed the signal by removing any outliers. Given the signal’s average (*µ*), the signal’s standard deviation (*σ*) and each observation *x*, an outlier was marked if *abs* (*x*) ≥ *µ* ± 6*σ*, and replaced by linear interpolation with its direct neighbors. The data was then z-scored to ensure for inter-subjects’ comparability. The data was detrended by subtracting a first-degree polynomial fitted using least squares. A Welch periodogram was then deployed with a window size of 512 samples (with an overlap of 128 samples and an NFFT of 4096), with samples collected every 10 minutes. The resulting power spectrum was normalized dividing by its total sum (i.e., 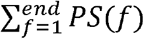). We then assessed in how many signals showed a peak around circadian, ultradian and multidien frequencies. Specifically, we focused on 24-hour periodicity (i.e., circadian); periodicity over multiple days (i.e., multidien) and periodicity within 24h (i.e., ultradian). The height of these peaks was computed to assess the strength of the periodic rhythm with that frequency.

### Responsiveness assessment

We divided patients into four classes based on their percentage of self-reported seizure reduction at their last follow-up compared to baseline (i.e., pre-implantation): (i) *super* (or level 3) responders (75-100%) (ii) *level 2 responders* (50-75%) (iii) *level 1 responders* (25-50%) and (iv) *non-responders* (<25%) responsiveness levels. In analogy to previous studies, class (i) and (ii) were merged as the “responders” group, while class (iii) and (iv) were classified as “non-responders”.

### Statistical and Data Analysis

#### Group comparisons

We first analyzed differences between clinical responsiveness that were *independent* of time evolution, focusing on two scenarios: (i) analysis of post-implantation measurements only to assess the presence of biomarkers able to predict therapeutic response in early stages and (ii) all collected data points over time, to identify if responsiveness could be related to a baseline shift in power spectra.

When comparing two levels of responsiveness (i.e., responders vs non-responders, or super-responders vs all others), we deployed a Mann-Whitney rank-sum U-test.

When considering four levels of responsiveness, we conducted a Spearman’s correlation test to test if any increasing or decreasing (not necessarily linear) trend could be found between the power spectrum data and the seizure frequency reduction. N-way ANOVA with post-hoc Tukey-HSD multiple comparison correction was applied to assess which groups were statistically different.

#### Time evolution

We assessed the temporal correlation between spectral band power and time (correcting for the effect of responsiveness) and deployed *Generalized Additive Mixed Models (GAMM)*, which allow to account for inter-thalamic variability (random variable) and to consider non-linear relationships between the outcome variable (PS) and the independent variable (time) by using *splines*. Splines are smooth, flexible functions made by connecting polynomial segments at certain points called knots. They allow to model non-linear relationships without pre-defining a default non-linear function (for instance, a quadratic or exponential function)^56^. The `mgcv` package was used in R Statistical Software (v 4.3.2, R Core Team 2021)^57^.

Specifically, we modeled a baseline group difference between responders and non-responders, separate time trends for each group, and thalamic-level random intercepts to account for individual variability.

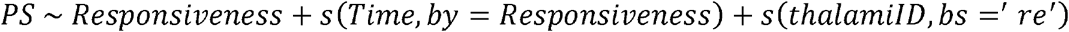

A model was fitted for each frequency band of interest. Here, PS is the spectral power, Responsiveness is a fixed effect (with either 2 or 4 levels of responsiveness), *“s”* indicates a spline (smooth curve) over time, which is fitted to the data for each of the two levels of responsiveness. The thalamus is modeled using a basis (*bs*) as a random effect (‘*re*’) that models thalamic-specific intercept.

Importantly, in this statistical model, we only considered data up to 2 years post-implantation due to the data imbalance between responders and non-responders (only 2 non-responders were followed-up afterwards).

If the model revealed significance in the second term (*s (Time, bγ = Responsiveness*)), we further calculated the pairwise difference between the predicted spline (i.e., evolution) for responders and non-responders at each time-point. Differences with non-overlapping confidence interval (CI) were considered significant. In the results section, we present the earliest time at which discrimination could be identified for each band and contact.

#### Circadian data

When comparing the peak height of circadian (24-hours) and ultradian (12-hours) rhythms in responders versus non-responders in the Chronic Sensing data, we used a Mann-Whitney rank-sum U-test.

Significance levels are reported as follows: * (p<0.05), ** (p<0.01), *** (p<0.001), **** (p<0.0001). Further details on which test was deployed for each result are reported in the Results section. The data analysis was performed using **MATLAB** (using the open-access toolbox *fieldtrip*^58^), **Python** (using the open-access toolbox *MNE* ^59^) and **R** (using the open-access library mgcv^57^).

## Results

A cohort of N = 22 epilepsy patients, implanted with bilateral DBS leads in the anterior nucleus of the thalamus (ANT) (Figure 1A), underwent ANT LFP recordings at multiple time points post-implantation either during outpatient visits (“*LFP Streaming*”) (*N*_*rec*_ = 104, in *N*_*subj*_ =20) or in long-term recordings performed in *Chronic Sensing Mode* (*N*_*rec*_ = 27, in *N*_*subj*_ =12) (Figure 1B-C).

We were able to follow up patients for a long observational period, with some patients reaching 4-years post-implantation follow-up visits (Figure 1D-E). The first recording (what we defined as post-implantation baseline) was performed at 138.15 ± 184.02 (mean ± standard deviation) days post-implantation, with a median of 63.5 days post implantation (two months). The last recording was performed at 621.81 ± 386.26 days post-implantation.

For the *N*_*subj*_ =20 with LFP streaming recordings, we analyzed the average spectral power (across follow-ups) in the three bipolar montages of interest (*V*_0_|_1_, *V*_1_|_2_ and *V*_2|3_). We observed that ANT activity exhibited a classical aperiodic 1/*f* component, with peaks at *δ, θ, α* and *β* visible across the three montages (Figure 1F,G) but not for all patients.

Based on their seizure frequency reduction at the last follow-up, patients were divided into 4 groups: super-responders (i.e., level 3 responders) (– Δ*SF*% = 75-100%, *N*_*subj*_ /*N*_*all*_=27%), level 2 responders (– Δ*SF*% = 50 – 75%, *N*_*subj*_ /*N*_*all*_=41%), level 1 responders ( Δ*SF*% = 25 – 50%, *N*_*subj*_ /*N*_*all*_=18%) and non-responders (– Δ*SF*% = 0 – 25%, *N*_*subj*_ /*N*_*all*_=14%) (Figure 1H). Further relevant information regarding patients’ demographic characteristics can be found in Table 1.

### ANT activity changes over time, concurrently with patient-specific therapeutic efficacy

We first analyzed whether LFP power in different predefined frequency bands was changing over time by averaging all observations collected from the cohort during the same time interval (Figure 2A). While not all patients were recorded at each interval of interest, we were able to gather data from at least 10 patients up to 3 years post-implantation, with only 3 patients receiving a follow-up in the interval of 3 to 4 years post-implantation (Figure 1H). For each thalamus, we normalized the power spectrum according to its first recorded value post-implantation *PS* (*t* = *t*_0_), to assess the evolution comparatively across subjects. The analysis was separately performed for *V*_0|1_, *V*_1|2_ and *V*_2|3_, but only contact *V*_1|2_ will be reported in the main figure, as it is centrally located and therefore fully within the ANT in all patients. Results from two other bipolar derivations are reported in the Supplementary Material (unless specified).

**Figure 2.**
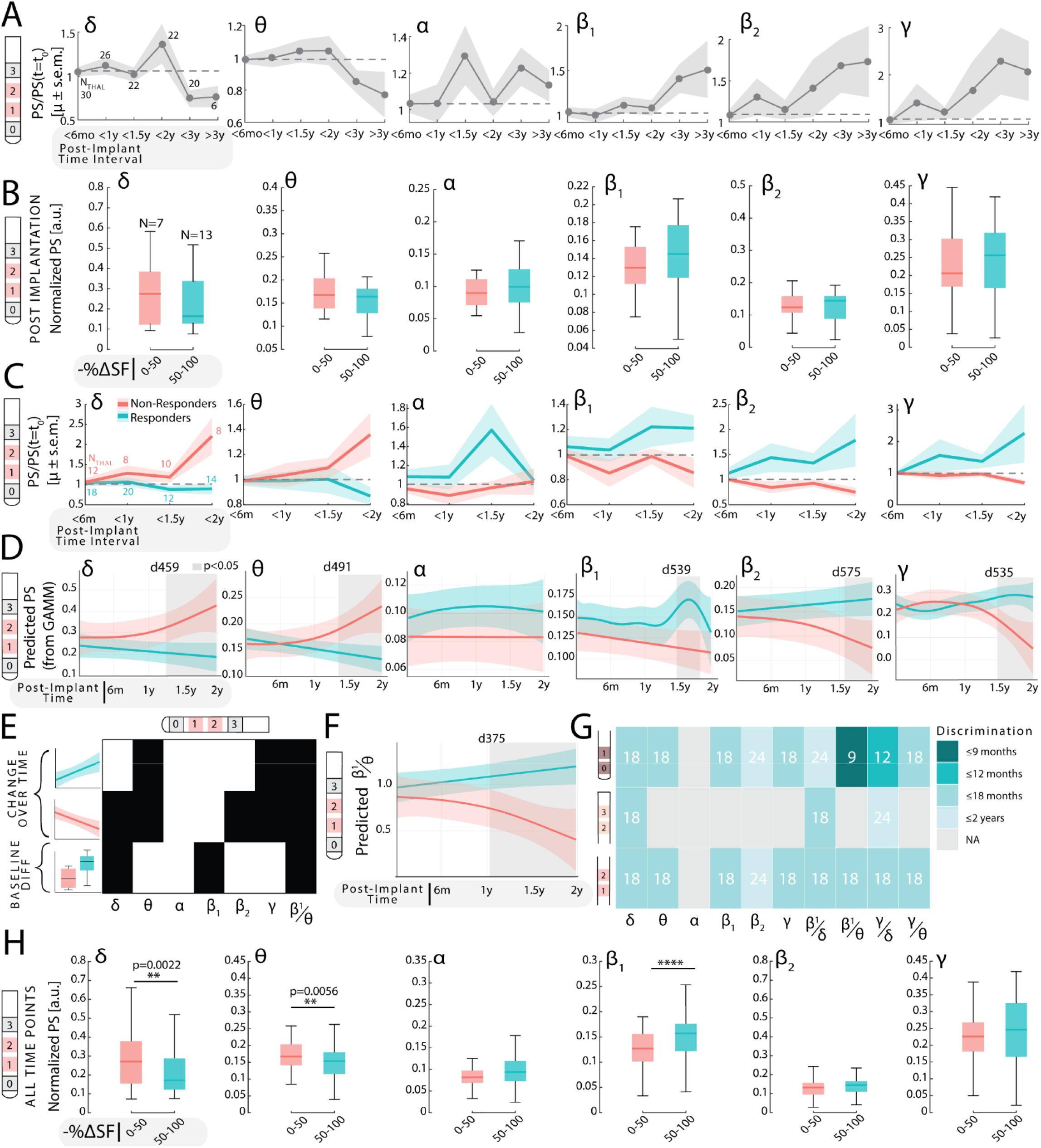
A) Evolution of the normalized PS for each band (mean ± s.e.m.) in contact pair 1-2. B) Post-implantation PS for each of the two classes, only for the montage 1-2. N= represents the number of subjects in each group. C) Raw evolution of the normalized PS for each band and each responsiveness group. Data was averaged for each time-interval (6-months long). *N*_*THAL*_ represents the number of thalami. D) Population-level predictions from the GAMM model for responders and non-responders. A grey box indicates significance (i.e., non-overlapping confidence-intervals). The earliest day of non-overlapping CI Is reported. E) For each fixed-effect of the model (evolution for responders, evolution for non-responders and baseline difference) and each band, significance as a predictor is reported (p<0.05). F) GAMM predictions for the ratio *β*_1_ /*θ*. A grey box indicates significance, and the earliest day of non-overlapping CI is reported. G) For each band and each contact pair, the earliest time for discrimination is reported in months post-implantation. H) Merged observations over-time for the two-groups with reported Mann-Whithney U-tests and statistical significance.

Interestingly, we have found a diverging temporal behavior between ANT LFP in lower frequencies (*δ, θ*), which showed a decreasing trend over time, and higher frequencies (*α, β*_1_, *β*_2_, *γ*) which increased over time (Figure 2A). While subtle differences were present, these trends were similarly expressed across the dorsoventral axis of the ANT (Supplementary Figure 2). However, the high-variability across thalami prevented the detection of a statistically significant effect of time on ANT LFP band power in the full cohort.

We therefore proceeded by assessing whether this variability could be attributed to the variance in therapeutic response, dividing our cohort in two groups based on a 50% seizure frequency reduction threshold (Responders (− Δ*SF*%≥ 50%)=13, Non-Responders(− Δ*SF*% ≤ 50%) =7), following the current state of the art definition of responsiveness to neuromodulation^6^. Patients with a seizure frequency reduction of less than 50% will hereafter be referred to as ‘non-responders’.

In the first measurement post-implantation, there were no significant differences between responders and non-responders in any band (Figure 2B, p<0.05, Mann-Whithney U-test), suggesting that DBS outcome could not be discriminated early on after surgical implantation. We then examined power changes in responders and non-responders over time, to understand how the evolution, rather than baseline, post-operative measurements, of the ANT spectral properties could be related to therapeutic efficacy. Here, we observed that responders and non-responders presented different evolutions over time, suggesting that continuous DBS modulates ANT dynamics in an outcome-specific way (Figure 2C).

We then used a Generalized Additive Mixed Model (GAMM) to model the non-linear relationships between time and PS. This model assessed the significance of time in predicting power spectrum changes when accounting for the responsiveness of the patients. We modeled the power spectral evolution as a function of responsiveness (with two possible levels), adding an offset in the interpolating spline, and the non-linear effect of time on power spectrum (by considering one spline for each responsiveness level, assuming responders and non-responders presented potentially independent dynamics). Inter-thalamic variability was accounted for by adding it as a random effect. As no poor responder was followed-up after 2-years post-implantation, we restricted this analysis up to this period. Using the GAMM, we found that PS changes significantly over-time, but most importantly, does so in a responsiveness-dependent manner. Given the model:

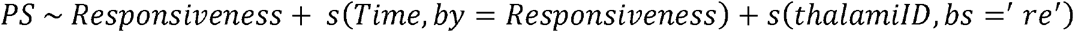

we hereby report the significance of the predictors for the contact pair 1-2.

We observed that responders and non-responders presented diverging behaviors over-time, as represented by the population-level predictions from the GAMM models in different frequency bands (Table 2, Figure 2D).

**Table 2.** GAMM (50% cut-off responsiveness): Results of the GAMM for each frequency band and contact 1-2. For the splines, effective degrees of freedom (representing the complexity of the spline, edf) and p-values are reported. The model fit is reported using adjusted R^2 and deviance. In the responsiveness fixed effect, “Pr(>|t|)” indicates the p-value from the regression-based t-statistic, and “Estimate” indicates the estimate of the linear fit (if positive, a higher PS is correlated to a better responsiveness). If the Responsiveness fixed effect is significant, the two levels of responsiveness show a significantly different power spectrum (independent on time). If the splines are reported as significant for a specific responsiveness group, that group has a significant evolution over time (i.e., ANT dynamics changes in a significant way).

| $V_{1\vert 2}$ | **Responsiveness (fixed effect): Responsiveness_1 - Responsiveness_0** | **$s(\text{Time}): \text{Responsiveness}_0$** | **$s(\text{Time}): \text{Responsiveness}_1$** | **Model Fit** |
| --- | --- | --- | --- | --- |
| $\delta$ | Estimate: -2.181<br>**Pr(>\|t\|): 0.0309 (\*)** | **P-value: 0.035 (\*)**<br>edf: 1.715 | P-value: 0.212<br>edf: 1.000 | $R^2_{adj} = 0.492$<br>Deviance = 58.7% |
| $\theta$ | Estimate: -0.017<br>Pr(>\|t\|) = 0.218 | **P-value: 0.00875 (\*\*)**<br>edf: 1.868 | **P-value: 0.01613 (\*)**<br>edf: 1.000 | $R^2_{adj} = 0.47$<br>Deviance = 56.4% |
| $\alpha$ | Estimate: 0.0186<br>Pr(>\|t\|) = 0.093 | P-value: 0.973<br>edf: 1.000 | P-value: 0.590<br>edf: 1.615 | $R^2_{adj} = 0.523$<br>Deviance = 61% |
| $\beta_1$ | Estimate: 0.0262<br>**Pr(>\|t\|) = 0.0259(\*)** | P-value: 0.123<br>edf: 1.000 | P-value: 0.314<br>edf: 5.584 | $R^2_{adj} = 0.47$<br>Deviance = 57.9% |
| $\beta_2$ | Estimate: 0.03506<br>Pr(>\|t\|) = 0.154 | **P-value: 0.0142(\*)**<br>edf: 1.622 | P-value: 0.1372<br>edf: 1.000 | $R^2_{adj} = 0.838$<br>Deviance = 87.4% |
| $\gamma$ | Estimate: 0.0235<br>Pr(>\|t\|) = 0.419 | **P-value: 7.5e-05 (\*\*\*)**<br>edf: 2.894 | **P-value: 0.0231 (\*)**<br>edf: 3.874 | $R^2_{adj} = 0.658$<br>Deviance = 73.8% |

**Table 3.** GAMM (Super-responders vs all): Results of the GAMM for each frequency band and contact 1-2. For the splines, effective degrees of freedom (representing the complexity of the spline, edf) and p-values are reported. The model fit is reported using adjusted R^2^ and deviance. In the responsiveness fixed effect, “Pr(>|t|)” indicates the p-value from the regression-based t-statistic, and “Estimate” indicates the estimate of the linear fit (if positive, a higher PS is correlated to a better responsiveness). If the Responsiveness fixed effect is significant, the two levels of responsiveness show a significantly different power spectrum (independent on time). If the splines are reported as significant for a specific responsiveness group, that group has a significant evolution over time (i.e., ANT dynamics changes in a significant way).

| $V_{1\vert 2}$ | *Responsiveness (fixed effect):*<br>*Responsiveness_Super*<br>*-Responsiveness_0* | *s(Time):Responsiveness_0* | *s(Time):Responsiveness_Super* | *Model Fit* |
| --- | --- | --- | --- | --- |
| $\delta$ | Estimate: -0.133<br>**Pr(>\|t\|): 8.25e-04 (\*\*\*)** | P-value: 0.2599<br>edf: 1.000 | P-value: 0.20<br>edf: 1.000 | $R^2_{adj} = 0.47$<br>Deviance = 56.4% |
| $\theta$ | Estimate: -0.022<br>Pr(>\|t\|) = 0.13 | P-value: 0.1544<br>edf: 1.000 | **P-value: 0.0035 (\*\*)**<br>**edf: 1.000** | $R^2_{adj} = 0.45$<br>Deviance = 53.9% |
| $\alpha$ | Estimate: 0.011<br>Pr(>\|t\|) = 0.298 | P-value: 0.56<br>edf: 1.000 | P-value: 0.74<br>edf: 1.000 | $R^2_{adj} = 0.52$<br>Deviance = 60.0% |
| $\beta_1$ | Estimate: 0.039<br>**Pr(>\|t\|) = 0.0015(\*\*)** | P-value: 0.42<br>edf: 1.35 | P-value: 0.28<br>edf: 1.000 | $R^2_{adj} = 0.46$<br>Deviance = 55.5% |
| $\beta_2$ | Estimate: -0.001<br>Pr(>\|t\|) = 0.96 | P-value: 0.13<br>edf: 2.02 | P-value: 0.088<br>edf: 1.000 | $R^2_{adj} = 0.83$<br>Deviance = 86.8% |
| $\gamma$ | Estimate: 0.10<br>Pr(>\|t\|) = **4.10e-04 (\*\*\*)** | **P-value: 0.11**<br>edf: 1.000 | **P-value: 0.004 (\*\*)**<br>**edf: 1.000** | $R^2_{adj} = 0.598$<br>Deviance = 67.8% |

Statistically-significant differences across the two groups were arising over-time, with a minimum time lag of discrimination of the two groups at 459 days post-implantation (*δ*) and a maximum of 575 days post-implantation (*β*_2_).

Analyzing the effect of time on the power spectral dynamics separately for each group, we found that non-responders showed a significant change in spectral power in *δ, θ, β*_2_ and *γ*. These effects were strongly non-linear, as showed by the estimated degrees of freedom (Table 2). Responders, on the other hand, presented significant changes in PS only for *θ* (linear decrease, edf=1.000) and *γ* (non-linear). Responsiveness alone had a (significant) fixed effect in *δ* and *β*_1_, meaning that there was a baseline difference between the two groups in these two frequency bands. For all frequency bands, the random effect of the thalamus (represented by the thalamic ID), was significant (p<2*e*^−16^), meaning that each thalamus has a non-negligible different baseline level in modeling the predictors-outcome relationship. Significant fixed effects of the model (p<0.05) are summarized graphically in Figure 2E. Interestingly, LFP power spectrum in *θ* was the only frequency band showing a significant change both in responders and in non-responders.

As lower frequencies (*δ, θ*) and high frequencies (≥ *α*) expressed opposite relationships to therapeutic efficacy, we formulated a high-to-low ratio between high and low band power to maximize the difference between responders and non-responders. The ratio in power spectrum between *β*_1_ and *θ* (*β*_1_ /*θ*) showed a significant fixed-effect of the responsiveness level (Estimate=0.29, Pr(>|t|)=0.0131and a significant effect of time for both responders and non-responders (s(Time):Responsiveness_0, edf=1.626, p-value: 0.00899; **s**(Time):Responsiveness_1, edf=1.000, p-value: 0.04628) (Figure 2E). Interestingly, when observing the population-level predictions of the GAMM of *β*_1_/*θ* over-time, differences between responders and non-responders arised as early as 375 days post-implantation (Figure 2F), almost 3 months earlier than when considering *δ* power only.

We then analyzed the earliest time point post-implantation at which discrimination between the two is present (defined by non-overlapping confidence intervals of the population-level GAMM predictions). Among the contact pairs, the uppermost bipolar montage (3-2) showed the weakest discriminatory power, while contacts 0-1 and 1-2 demonstrated the ability to discriminate between groups, with a minimum discernible difference observed at 18 months post-implantation. However, when examining high-to-low frequency ratios, the minimum discrimination time was reduced by half, with contact 0-1 allowing for group differentiation as early as 9 months post-implantation (Figure 2G).

When merging all the collected samples over-time, and comparing the two groups (Mann-Whithney U-test, p<0.0083 for significance with Bonferroni multiple comparison correction), we found significant differences among the two groups in *δ, θ* and *β*_1_ (p-values: *δ*: 0.002, θ: 0.006, *β*_1_ : 2.6e^−04^, Figure 2H).

### ANT dynamics predicts super-response to therapy by month 1 post-implantation

Building on the binary classification (≤50% vs >50% response), we next investigated the temporal dynamics of ANT-LFP in a more granular analysis using the four-level responsiveness scale as defined above.

In post-implantation measurements, we found that super-responders (level 3) had the lowest *δ, θ* LFP power, and the highest power in *α, β*_1_, *β*_2_ and *γ* frequency bands. By deploying Spearman’s correlation, we assessed the presence of a positive relationship between the PS (calculated from *V*_1|2_) at post-implantation and the efficacy of stimulation. We noticed that *δ* and *γ* power were significantly lower in super-responders compared to the three remaining levels of responsiveness (Figure 3A), when comparing across the four levels of responsiveness with ANOVA-N (and post-hoc Tukey-HSD). This implies that super-responders show specific neurophysiological properties already at baseline measurements.

**Figure 3.**
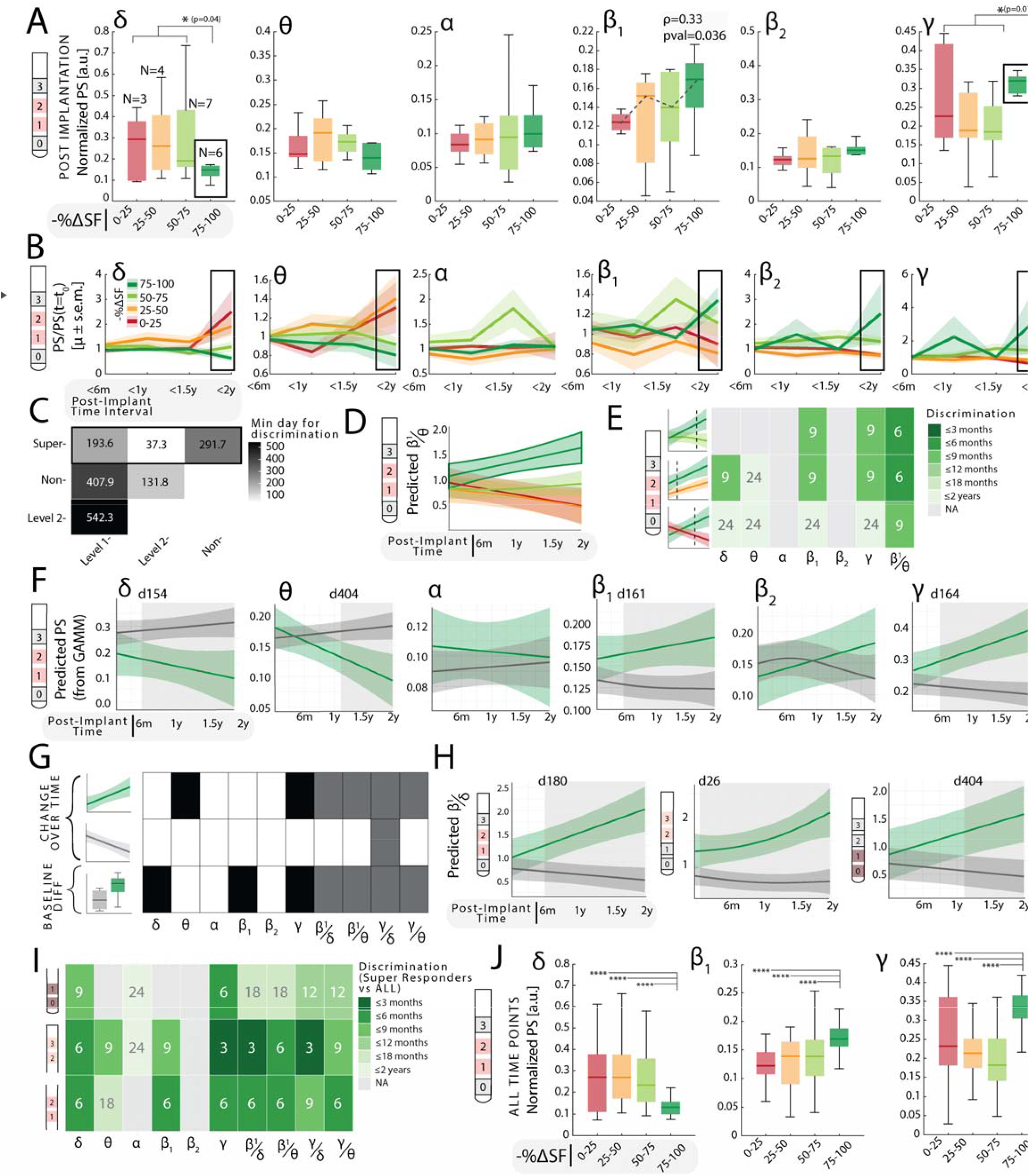
A) Post-implantation PS for each of the four classes, only for the montage 1-2. N= represents the number of subjects in each group. A dashed line in *β*_1_ indicates a significant Spearman’s correlation (p and p-val are reported). Post-hoc Tukey-HSD results are reported. B) Raw evolution of the normalized PS for each band and each responsiveness group. Data was averaged for each time-interval (6-months long). C) The minimum time of discrimination across the four classes is reported as days post-implantation. A black empty box indicates the class with the lowest time for discrimination. D) GAMM predictions for the ratio *β*_1_ /*θ* for the four levels. E) For contact pair 1-2, the earliest time for discrimination is reported in months post-implantation for super-responders vs each of the other groups. F) Population-level predictions from the GAMM model for super-responders vs sub-optimal responders. A grey box indicates significance (i.e., non-overlapping confidence-intervals). The earliest day of non-overlapping CI Is reported. G) For each fixed-effect of the model (evolution for super-responders, evolution for suboptimal-responders and baseline difference) and each band, significance as a predictor is reported (p<0.05). H) GAMM predictions for the ratio *β*_1_ /*δ*. A grey box indicates significance, and the earliest day of non-overlapping CI is reported. I) For each band and each contact pair, the earliest time for discrimination is reported in months post-implantation. J) Merged observations over-time for the four-groups with reported ANOVA-N with post-hoc Tukey-HSD and statistical significance.

Following the same analytical approach as above in the dichotomous analysis, we examined the evolution of the LFP power spectrum over time, grouping observations into predefined 6-month intervals. Level 2 responders and super-responders (light and dark green, respectively) showed progressively diverging patterns over time when compared to level 1 and non-responders (orange and red), as emphasized by the black box in the panel (Figure 3B). This reinforces the idea that DBS shapes ANT-LFP dynamics in diverging ways for responders and non-responders.

To determine the earliest time point at which responders’ group could be differentiated, we examined the population-level predictions generated by the GAMM, focusing on the first time interval where confidence intervals no longer overlapped (Figure 3C). We found that super-responders could be discriminated earlier than other groups by identifying the minimum time of discrimination across all contacts and frequency bands. Additionally, when analyzing high-to-low frequency ratios (e.g., *β*_1_ /*θ*), super-responders again displayed distinct early dynamics compared to the other three response levels (Figure 3D). Focusing on LFP data from the contact pair 1–2, we found that super-responders could be differentiated from level 1 and level 2 responders as early as 6 months post-implantation, based on the *β*_1_ /*θ* ratio. Discrimination from non-responders was achieved slightly later, at 9 months post-implantation (Figure 3E).

To further contrast optimal versus sub-optimal clinical trajectories, we grouped level 1 and level 2 responders, and non-responders into a single “sub-optimal outcome” group (N = 14), and compared them against the super-responders (N = 6). Strikingly, population-level GAMM predictions revealed early divergence in trajectories, with *δ* power showing significant separation as early as 154 days post-implantation (Figure 3F). Here, all bands except *α* and *β*_2_ showed progressive divergence in time.

Super-responders exhibited a significant linear (as shown by edf = 1.000) decrease in *θ* power and a linear increase in *γ* power over time, reinforcing prior findings (Table 1). This suggests that a reduction in *θ* power over time and an increase in *γ* power (compared to baseline) is a prognostically good indicator for treatment response. Baseline differences between groups were evident in *δ, β*_1_ and *γ* power. Incorporating high-to-low frequency ratios further enhanced discriminatory power, revealing both distinct baseline values and temporal dynamics in super-responders across all proposed ratios. Among sub-optimal outcome patients, non-responders showed a significant evolution only in the *γ*/*δ* ratio (Figure 3G).

We report as an example the evolution of *β*_1_ /*δ* across the three contact pairs. Here, discrimination across the two groups was possible as early as 26 days post-implantation in contact 2-3 (Figure 3H).

Among all contact pairs, the most superior contact pair exhibited the earliest discrimination between groups— occurring before three months post-implantation—while the inferior contacts revealed later discrimination, with separation in the *β*_1_/*δ* ratio emerging only after 18 months. (Figure 3I).

When aggregating data across the full follow-up period, the trends observed post-implantation became more pronounced. Super-responders demonstrated significant and robust differences (p < 0.0001) from all other groups in *δ, β*_1_, and *γ* power (Figure 3J), confirming the stability and strength of these spectral biomarkers over time.

### ANT activity shows relevant circadian and multidien rhythms, which may be relevant for therapeutic outcomes

Finally, we analyzed long-term *Chronic Sensing* recordings. As these measurements in *N*_*subj*_ =12 were retrospectively investigated, each frequency was chosen on an individual level, and the recorded frequency bands were not evenly distributed (Figure 4A). We mostly recorded data in the *θ* band (*N*_*samples*_=33), followed by *γ* (*N*_*samples*_=10), *α* (*N*_*samples*_=5) and *β*_1_ (*N*_*samples*_=2).

**Figure 4.**
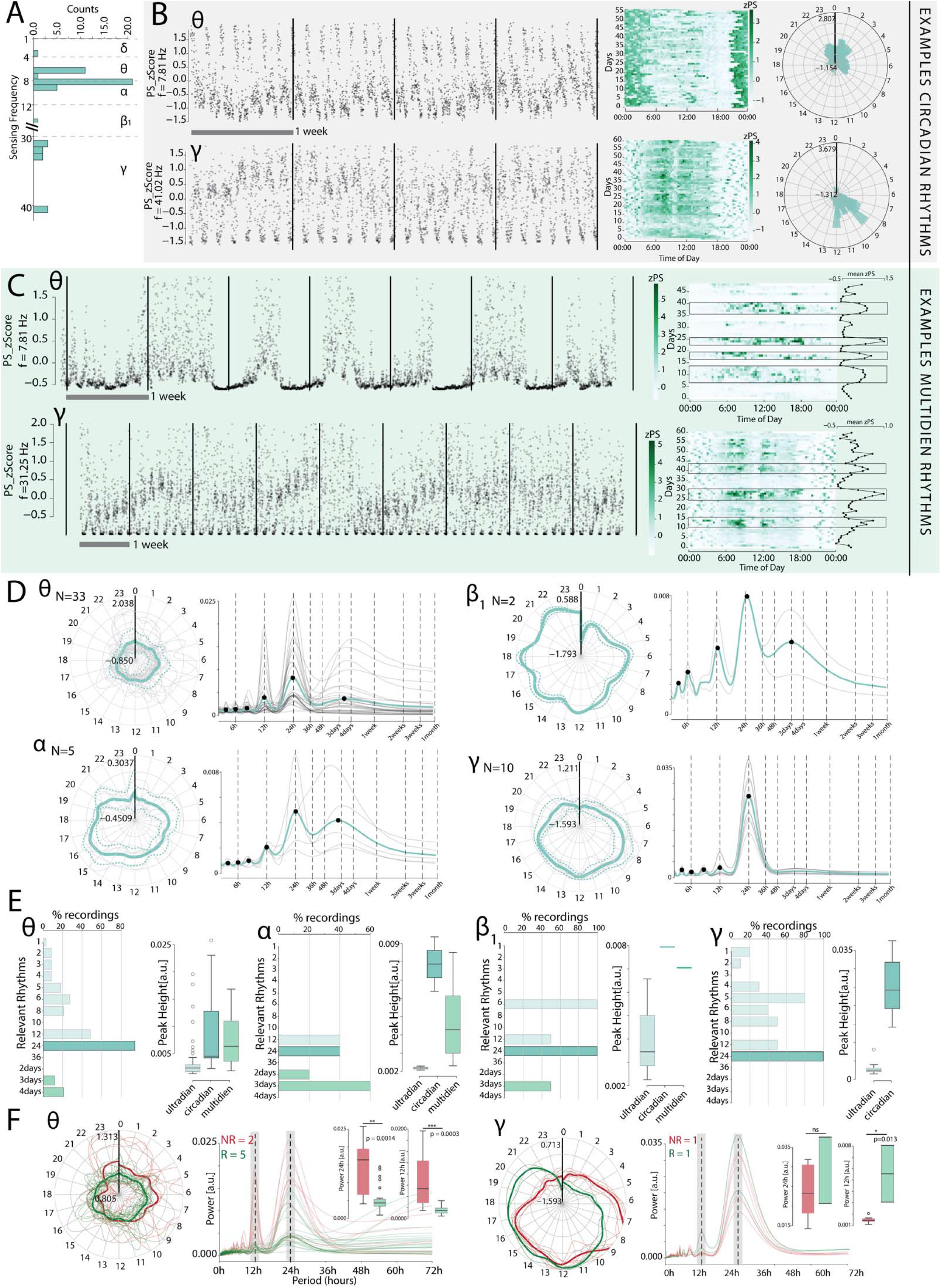
A) Sensing frequencies distribution B) Two patients raw LFP power over a month (left), day-time of day distribution of power (middle) and polar histogram over time of the day (right). C) For two patients, raw LFP is reported to show multidien rhythms as in panel B. The day-time of day distribution is also reported, together with the average PS over days (left inlet). D) Left: polar distribution of the band power over time of the day. The thick, blue line indicates the mean value, the dashed blue lines indicate the standard error of the mean, and the grey lines indicate each recording separately. Right: Welch’s periodogram of the relevant rhythms. The thick blue line indicates the mean, and the grey lines indicate each periodogram separately. Detected peak times are reported as black dots. E) For each band, the % of recordings showing ultradian, circadian and multidien rhythms is reported for each relevant frequency. The peak height at each of the three groups is also shown. F) For *θ* and *γ* bands, the same results are reported separately for responders (i.e., class (i) and (ii)) and non-responders (i.e., class (iii) and class (iv)), together with the population peak height at 24 and 12 hours and their statistical significance (Mann-Whitney test, p<0.05). E)

First we observed the ANT LFP activity is governed by circadian rhythms (i.e., 24-hour periodicity) although there was also highly individual pattern of longer-period (multidien) cyclical activity. We report two patients’ examples that are depicted in Figure 4B (left side), where the patient-specific evolution of *θ* and *γ* is shown over a month (with one observation every 10 minutes). While in both patients a 24-hour power periodicity is present, *θ* activity in one subject was highly emphasized in night-time, and *γ* activity in the second subject was mostly present during day hours (Figure 4B, middle and right part). Similarly, we also found individual multidien cyclical activity. We show two subjects’ ANT activity (Figure 4C) in the same bands. While the underlying circadian rhythm is preserved, an additional oscillatory component exists where the ANT power at *θ* and *γ* is modulated over days with baseline shifts.

We have then considered each of the existing observations for the analyzed bands to understand if the distribution of the power over time of the day (left side of the plot) and the underlying rhythms (right side of the plot) were consistently present across different patients. We found that *α, β*_1_ and *γ* power were mostly present during daytime, while *θ* distribution was less concentrated in a specific moment of the day and more spread throughout day and night-time (Figure 4D).

We then counted the number of samples with a peak in the periodogram at frequencies of interest (i.e., < 24 hours, 24 hours, >24 hours) and the peak height (intended as the power of that oscillation) in circadian (24-hours), ultradian (<24 hours) and multidien (>24 hours) groups. The majority of observations in the *θ* band presented a circadian rhythm at 24-hours (Figure 4E), followed in prevalence by a peak at 12 hours. 20% of the recordings presented a multidien peak at 4 days. Also, multidien and circadian rhythms had similar power, with ultradian rhythms being associated less strongly to *θ* oscillations. In *α* band, peaks at 12, 24, 2 and 3 days were visible, with the highest power in circadian oscillations. In the *β*_1_ band, both observations had peak at 24 and 6 hours. Finally, when analyzing *γ*-band activity, no multidien rhythm was detected but all of the observations presented a circadian rhythm (Figure 4E).

Finally, we assessed whether individual rhythmicity could be related to DBS response. Intriguingly, in 2 non-responders and 5 responders with *θ*-band recordings, we observed that non-responders had a stronger modulation of *θ* oscillations along the circadian rhythm.

## Discussion

Anterior thalamic (ANT) DBS is a promising approach for the treatment for patients with drug refractory epilepsy but only results in a median 50-65% seizure reduction rate (at 5-years post-implantation). Among other factors that limit the understanding of this clinical outcome variability, two main challenges lie in the post-implantation therapeutic strategy. Firstly, no neurophysiological biomarker for the prediction and monitoring of therapeutic efficacy has been defined yet. Secondly, *adaptive* DBS, which relies on intrinsic neural rhythms and states to guide patient-specific stimulation protocols, is currently not used in epilepsy patients due to a lack of understanding on how to clearly interpret ANT neurophysiological dynamics. Therefore, identifying reliable biomarkers within ANT activity that reflect long-term therapeutic outcomes and incorporating patient-specific physiological rhythms, such as circadian fluctuations is key to further optimize therapeutic success and treatment strategy.

Here we analyzed a large monocentric longitudinal dataset with chronic and intermittent intracranial thalamic LFP recordings over a follow-up period of up to four years in an outpatient setting. As a core finding of the study, we identified distinct patterns of ANT LFP spectral properties that differentiate DBS responders from non-responders at baseline post-implantation and later in the temporal evolution of LFP biomarkers. Furthermore, we found evidence that spectral temporal dynamics within the ANT evolve in a manner that is specific to optimal clinical outcome (DBS super-responders), and that these changes can be detected before seizure-diary based assessments. Our findings also support the notion that ANT-DBS acts via chronic neuromodulation of thalamocortical circuits, and that this process involves detectable spectral temporal dynamics in an outcome-specific fashion. Furthermore, we reveal novel insights into the physiological rhythmicity of ANT activity and its potential relevance for optimizing therapeutic strategy.

These findings together hold promise for enabling earlier, physiology-guided calibration of stimulation protocols in a therapy that is otherwise characterized by protracted and empirical optimization.

### ANT activity holds predictive biomarkers of therapeutic efficacy

One major finding of this study is that post-implantation ANT oscillations evolve in specific temporal trajectories based on clinical response, with responders showing a general increase in higher-frequency activity (especially *β*_1_ and *γ* bands), and a decrease in lower frequency bands (*δ* and *θ*), whereas non-responders exhibited an opposite dynamic, possibly suggesting maladaptive changes in oscillatory activity.

We initially divided our cohorts based on a 50% cut-off threshold in seizure frequency reduction, following default clinical definitions on responsiveness to neuromodulation. Here, no significant spectral differences could be observed at baseline (post-implantation measurements) across the two groups, but the longitudinal evolution of their power spectra revealed robust divergences, suggesting that it is not the initial state alone, but the direction of spectral change over time that carries predictive information about therapeutic success. This notion is particularly significant in light of the chronic and slowly emerging clinical effects of ANT-DBS, which are believed to be mediated by long-term network reorganization and plasticity. The gradual differentiation of spectral profiles over time aligns with this hypothesis, reinforcing the idea that successful neuromodulation induces specific neurophysiological network changes that reflect underlying structural or functional adaptation^13^. Specifically, we observed that responders showed a significant linear decrease in *θ* band activity and a non-linear change in *γ* power.

The importance of *θ* oscillations in describing the diverging evolution of responders and non-responders ANT dynamics resonates with our previous findings where high ANT *θ* band activity during DBS electrode implantation was found to be predictive of good therapeutic outcome^55^. Recently, we also established that ANT *θ* band activity correlates with seizure likelihood in a SEEG study during presurgical evaluation. ^55^ Thus, higher *θ* band activity in the ANT might both represent a pathological biomarker for epileptic networks and correlate with high seizure burden. Thus, *θ* band activity could reflect an interesting neurophysiological proxy for the identification of the optimal stimulation site and the seizure suppressing network effect. In this study, we observed a marked *reduction* in *θ* oscillations after implantation and a continuous reduction in the temporal long-term evolution in clinical responders. This strongly supports the hypothesis that *θ* activity might reflect a pathological epilepsy biomarker that can be targeted, modulated and reduced by chronic ANT-DBS.

Group differences among the two cohorts could also be identified in *δ* and *β*_1_ frequency bands validating our previous findings where these bands were fundamental in discriminating between epileptogenic and non-epileptogenic ANT-based networks^55^. Indeed, the importance of *β*_1_ oscillations was also relevant in a recent pre-operative study, where SEEG recordings revealed that the activity of ANT-networks which were encompassing the seizure onset zone were orchestrated by *β*_1_ oscillations ^55^. Moreover during DBS implantation (pre stimulation), *β*_1_ activity was lower in responders compared to non-responders^50^. In the current study, the first post-implantation measurement showed *high β* activity in responders, increasing over time. Conversely to the role of *θ* oscillations, one could argue that *β*_1_ activity represents a physiological network that is strengthened over time through ANT-DBS. The diverging dynamics between lower frequencies (*δ, θ*) and higher frequencies (≥ *α*) in both responders and non-responders validated our previous results where we correlated ANT power spectrum to time to the next seizure^55^. This dichotomy suggested that the discrimination of therapeutic efficacy could be maximized by combining higher and lower frequencies. For this reason, we introduced high-to-low frequency ratios (e.g., *β*_1_/*θ*), which increased the predictive power of the model.

The predictive value of these biomarkers has potential translational implications to complement seizure diaries, which in some cases are only partially reliable. Upon validation of their correlation to seizure frequency evolution (and not only to outcome at last follow-up), ANT-LFP biomarkers may offer an objective and earlier estimate of treatment efficacy, potentially allowing for more timely adjustments in stimulation parameters or clinical strategy.

Although a 50% seizure reduction threshold remains a widely accepted clinical benchmark for defining responsiveness^6^, it may be insufficient to capture the heterogeneity in therapeutic outcomes. Patients above this threshold can indeed still differ markedly in long-term clinical outcome and physiological signatures. Moreover, achieving a “super-response”, i.e., a high seizure frequency reduction, is the clinical outcome that truly impacts quality of life. Identifying patients who show signs of ‘super-responsiveness’ or deviate from this trajectory may provide means to enable therapy optimization before the standard clinical re-assessment window. When we stratified patients using a four-level responsiveness scale, we were able to isolate the “super-responder” from all other patients. These patients, indeed, were not only characterized by a stronger clinical benefit (75–100% seizure reduction), but also presented a distinctive neurophysiological signature already at post-implantation measurements (*δ, γ*).

In follow-up recordings, the evolution of super-responders ANT activity could be discriminated earlier on from each of the remaining three groups. Specifically, differences arose early on in *δ, β*_1_ and *γ* (around 5 months post-implantation). Also in this analysis, the high-to-low (e.g., *β*_1_/*δ*) power ratio emerged as a particularly robust marker, showing both significant group-level effects and anticipating the earliest divergence between super-responders and the rest of the cohort than when considering single bands separately, finding a significant difference as early as 26 days post-implantation.

The concept of a high-to-low frequency ratio integrates both the enhancement of faster oscillations (potentially linked to cortico-thalamic engagement and plasticity) and the attenuation of slower rhythms (often associated with pathological synchrony), offering a succinct yet informative index of prediction and therapeutic progression. It also considers inter-individual differences in absolute power values by using an inherent within-subject normalization. Moreover, its relevance as a significant predictor both for baseline differences and fo outcome dynamics reinforces its potential role as both an early, post-implantation predictive marker and one to monitor longitudinally to gain insights into therapeutic efficacy.

Another interesting observation of the study lies in the heterogeneity of thalamic signal baselines across patients in both models (50% or 75% cut-off threshold), as captured by the significant random effect of thalamic ID in our models. This highlights the necessity of adopting patient-specific benchmarks rather than relying on fixed thresholds for biomarker interpretation. Furthermore, these findings pave the way for the potential value of closed-loop or adaptive DBS systems, which have the capacity to personalize stimulation based on evolving individual neurophysiology as opposed to static population norms.

### ANT Rhythmicity: Circadian and Multidien Oscillations

The temporal analysis of extended recordings revealed that ANT activity adheres to ultradian, circadian, and multidien rhythms. Some of these rhythms have been observed to be uniform across all patients, while simultaneously exhibiting individual variations. Circadian rhythms (24-hour periodicity) were predominant across all frequency bands. Interestingly, *α, β*_1_, and *γ* activity, associated with arousal and cognitive functions, was concentrated during daytime, while *θ* exhibited a less consistent pattern with some patients having nocturnal and some diurnal concentration. The presence of multidien rhythms adds a novel layer to our understanding of ANT activity. These oscillations may be indicative of underlying homeostatic or adaptive processes that link ANT signals and seizure activity. Given the fact that multidien seizure rhythms have been observed in long-term studies in humans, the subsequent logical step is to integrate these fluctuations with seizure timestamps and seizure clusters. Indeed, if a correlation was found among these two, the ANT activity could serve as a crucial biomarker of pro-ictal states to infer seizure risk.

Importantly, our results also suggest that stronger circadian rhythms in θ power were more prevalent in non-responders, while one responder exhibited a greater relative strength of ultradian (12-hour) rhythms. These findings must be interpreted with caution due to the small sample size and the lack of seizure data, but they raise the intriguing possibility that individual rhythmicity profiles could influence therapeutic outcomes. It is important to emphasize that the same subject could have received multiple Chronic Sensing recordings over the time-course of the follow-up. Therefore, it remains to be elucidated whether circadian rhythms might also influence the predictive outcome measures. The integration of seizure cycles could elucidate these mechanisms further. For instance, if *θ*-band circadian fluctuations correlated to seizure occurrence, and seizures also were subject to a circadian rhythmicity, a loss of such 24-hour rhythms could be beneficial. While these arguments remain hypothetical, the confirmation of the presence of physiological rhythms in the ANT is a crucial finding, as it could be leveraged for personalized stimulation protocols, paving the way for state-informed stimulation.

### Limitations and Future Directions

The present study is subject to several limitations. The most notable limitation is the relatively small sample size for long-term outpatient recordings (yet, the largest of its kind), particularly among non-responders, which limits the generalizability of the findings. The follow-up numbers exhibited a decline over time, with no patients in the 0–25% response group beyond three years. Furthermore, inconsistent follow-up timing necessitated the implementation of model-based interpolation, thereby introducing complexity to the assessment of continuous trends. While the therapeutic effects are anticipated to manifest over extended periods, the identification of short-term biomarker changes has the potential to facilitate more expeditious treatment decisions.

Patients were grouped based on their final responsiveness level, rather than correlating biomarkers with seizure frequency at each time point—an approach that could strengthen short-term predictive value. Furthermore, the variability in stimulation parameters (particularly amplitude and active contact location) complicates interpretation and necessitates further investigation into their influence on ANT dynamics.

Chronic sensing was limited to 60-day windows, restricting the ability to analyze longer-period cycles, such as multidien rhythms relevant for seizure clustering. Future studies should incorporate longer recordings, standardized follow-up protocols, and larger cohorts.

Finally, while strong correlations were observed between ANT activity and clinical response, the question of causality remains to be resolved. The integration of seizure and rhythm data, in conjunction with acute stimulation studies, could enhance our understanding of pro-ictal states and improve stimulation strategies.

Clinical Implication and Conclusion

Our findings demonstrate that ANT activity carries robust, time-evolving spectral biomarkers with significant clinical implications. By moving beyond cross-sectional analyses and embracing the longitudinal complexity of neural dynamic**s**, we have demonstrated that spectral biomarkers—especially when framed as evolving features—can help distinguish therapeutic outcomes earlier in the course of treatment.

The role of these biomarkers could be explored in relation to treatment strategy decisions - such as the calibration of stimulation parameters and the development of closed-loop applications with careful consideration. Based on our findings, future approaches could include adaptive stimulation parameters targeting a specific reduction in certain frequency bands. These biomarkers could thus provide clinicians with an objective tool to guide DBS optimization— complementing on seizure diaries and moving beyond empirical parameter tuning. Their predictive value within the first-year post-implantation supports a shift toward earlier, neurophysiology-informed adjustments to stimulation settings, replacing the current trial-and-error approach with more targeted and timely interventions.

Furthermore, the existence of circadian and multidien rhythms in ANT activity, particularly in relation to clinical response, emphasizes the value of incorporating temporal dynamics into stimulation protocols. These results provide a foundation for the development of adaptive, state-informed DBS strategies in epilepsy^24^, where stimulation is tailored not only to patient physiology but also to intrinsic neural timing.

Future work should validate these findings in larger and independent cohorts, explore the mechanistic underpinnings of these spectral changes, and link these biomarkers to seizure rhythms. In conclusion, these findings offer a framework for integrating objective neural signatures into DBS programming and highlight the potential for adaptive, patient-specific stimulation strategies. By embracing the dynamic nature of thalamic neuromodulation, this work opens new possibilities toward precision neurostimulation in epilepsy care.

## Data Availability

Informed consent was obtained from all patients or their legal representatives in compliance with the Declaration of Helsinki, and the study was approved by the local ethics committee (Kantonale Ethikkommission, KEK Zurich). However, this consent did not include provisions for making individual data publicly available.

## Acknowledgments

The authors are deeply grateful to the patients who participated in this study and consented to collect their data for the advancement of knowledge and for a better future for epilepsy patients. The authors would like to thank Dr Kai Lutz and Dr Gaetano Leogrande (Medtronic) for sharing technical information on the system. Medtronic had no impact on the study design, data acquisition or writing of the manuscript. The funder had no role in the experimental design, analysis, manuscript preparation, or submission. All authors had complete access to data. All authors authorized the submission of the manuscript.

This work was funded by a grant awarded to R.P. and L.I. from the Swiss National Science Foundation (SNF 197766). The authors report no competing interests.

